# ADVANCING ANTIPSYCHOTIC THERAPY: CLINICAL AND FORMULATION BENEFITS OF A NOVEL QUETIAPINE ORAL SUSPENSION FORMULATION

**DOI:** 10.1101/2025.09.02.25334902

**Authors:** Victoria Raboso, Katia Urso, Javier Gonzalez, Eva Garcia-Aguilar

**Author notes:** **Corresponding author:** Gonzalez, Javier (Medical Department; ITF Research Pharma, S.L.U.; Madrid, Spain).

## Abstract

**Background:** Quetiapine is an antipsychotic drug with unique pharmacological properties approved for the treatment of schizophrenia, bipolar disorder, and augmentation for major depressive disorder. In routine clinical practice, it is also frequently prescribed for conditions such as insomnia, anxiety, and psychomotor agitation.

Italfarmaco (ITF) Group has developed an innovative oral suspension formulation of quetiapine; its composition, optimized in viscosity and stability, aims to facilitate administration and dosing across a wide range of patient profiles, particularly those requiring low doses or those with difficulty swallowing, limited manual dexterity, or cognitive impairment.

**Objective:** This study aimed to evaluate the pharmacokinetic (PK) profile of this innovative oral suspension formulation of quetiapine compared to the immediate-release tablets.

**Methods:** A single-dose, open-label, randomized, two-sequence, two-treatment, two-period cross-over pivotal study was conducted in 43 healthy subjects under fasting conditions. Volunteers received a single 25 mg dose of quetiapine, administered either as an oral suspension or as an immediate-release film-coated tablet, with an appropriate washout between treatments. The PK performance of the two formulations was compared, maximum plasma concentration (C_max_), time to reach C_max_ (t_max_), area under the curve (AUC), and overall bioavailability. Plasmatic quetiapine levels were measured by validated high performance liquid chromatography (HPLC) methods blinded to the dosing randomization scheme.

**Results:** Quetiapine oral suspension demonstrated pharmacokinetic bioequivalence to the reference tablet in terms of C_max_ and AUC. It exhibited faster absorption, being detectable in plasma within 10 minutes following oral administration, with a median t_max_ of 0.75 h compared to 1.02 h for the immediate-release tablet. Calculating the median t_max_ difference for each subject shows that quetiapine oral suspension reaches peak plasma concentration about 30 minutes faster than the immediate-release tablet.

**Conclusion:** This novel quetiapine oral suspension formulation addresses key unmet needs in psychiatric pharmacotherapy treatment by combining a favorable pharmacokinetic profile with a patient-centered design. It enables precise dose titration of the drug and promotes faster absorption, thereby potentially achieving a more rapid onset of action. This formulation facilitates administration and dosing flexibility, an advantage for all patients, especially for the ones requiring low doses.

**KEY POINTS:**

- Italfarmaco (ITF) has developed an innovative oral suspension formulation of quetiapine.
- This novel quetiapine oral suspension formulation enables precise dose titration of the drug and promotes faster absorption, thereby potentially achieving a more rapid onset of action.
- ITF quetiapine suspension formulation is intended to address key unmet needs in antipsychotic pharmacotherapy treatment by combining a favorable pharmacokinetic profile with a patient-centered design.

## INTRODUCTION

Quetiapine is an atypical, second-generation antipsychotic drug, with unique pharmacological properties. Along with its active metabolite, norquetiapine, both quetiapine and norquetiapine display affinities to brain serotonin 5-HT_2A_, and dopamine D_1_ and D_2_ receptors, which are believed to contribute to its clinical antipsychotic efficacy, maintaining a low risk for extrapyramidal side effects [1-2]. See Table 1 for an overview of the pharmacodynamic profile of quetiapine [2].

**Table 1:** quetiapine activity in the central nervous system.

| **Doses** | **Low**<br>**(25-100 mg daily)** |  | **Medium**<br>**(300-600 mg daily)** | **High**<br>**(>800 mg daily)** |
| --- | --- | --- | --- | --- |
| **Receptors** | Histamine 1 | Alpha 1/ alpha 2<br>adrenergic | 5-HT1A, 5-HT2A/B,<br>5-HT2C | D2 |
| **Activity** | Antihistaminergic | Antiadrenergic | Serotonergic | Antidopaminergic |
| **Effect** | Sedation |  | Antidepressant properties | Antipsychotic properties |

Quetiapine is approved by United States and European regulatory agencies for the treatment of schizophrenia, bipolar disorder, and as augmentation treatment for major depressive disorder at a dose range of 25-800 mg/day. Also, and probably due to the sedative effect caused by H_1_ histamine receptor antagonism of quetiapine, and anxiolytic effect due to norepinephrine reuptake inhibition and serotonergic 5-HT_1A_ agonism of norquetiapine, quetiapine is extensively used at lower doses on routine clinical practice in insomnia, anxiety, and psychomotor agitation [1-3].

The management of psychiatric disorders frequently necessitates long-term pharmacological treatment, where adherence, tolerability, and individualized dosing are critical to achieve therapeutic success. Among second-generation antipsychotics, quetiapine is widely prescribed due to its broad efficacy and favorable tolerability profile. However, conventional solid oral formulations may not always meet the diverse needs of the patients, particularly in real-world settings where factors such as age, comorbidities, polypharmacy, and functional or cognitive limitations can affect treatment adherence and effectiveness.

Challenges such as difficulty swallowing, reduced dexterity, or cognitive impairment -common in elderly or institutionalized populations-can hinder the consistent use of tablet-based medications. These limitations underscore the need for more flexible and patient-friendly formulations that support individualized care [4-5].

To address these challenges, the Italfarmaco Group has developed an innovative oral suspension of quetiapine 25 mg per ml (patent pending), which has been evaluated and approved in the European Union through a Decentralized Procedure (DCP). This formulation is registered under the brand names Ketyalix^®^, Aquetia^®^, and Akelya^®^, among others. Designed with patient-centricity in mind, this quetiapine oral suspension is formulated at a concentration of 25 mg/mL, allowing for precise and flexible dose adjustments -from as low as 12.5 mg per dose up to 150– 200 mg-making it particularly advantageous for patients on low-dose regimens.

Italfarmaco quetiapine oral suspension incorporates several features that enhance usability and adherence. A carefully selected viscosity range minimizes the risk of laryngeal penetration or aspiration, contributing to an optimal safety profile during administration [6]. Moreover, its strawberry flavor, and the ability to be mixed with food, improves palatability and compliance, especially in geriatric or institutionalized populations. The excipient-minimized composition (free from sugars, lactose, gluten and with negligible sodium content) [7] supports its use in patients with dietary restrictions or polypharmacy concerns.

Finally, the product does not require refrigeration [7], simplifying storage and administration in both homecare and clinical settings.

These attributes align with the broader clinical benefits of liquid antipsychotic formulations, which can improve treatment adherence, reduce non-compliance, and facilitate safer titration protocols [1]. As such, this novel quetiapine oral suspension not only addresses a critical gap in the formulation landscape but also exemplifies the therapeutic potential of liquid antipsychotic delivery systems in modern psychiatric care.

The purpose of this study is to test the pharmacokinetic (PK) performance of this newly developed quetiapine oral suspension and compare it with the reference immediate-release film-coated tablet formulation, according to the European Medicines Agency (EMA)’s guideline on the investigation of bioequivalence recommendations.

## METHODS

A 25 mg/ml quetiapine oral suspension was developed by the Italfarmaco Group to ensure stability in different climatic conditions and to facilitate ease of handling and dosing (patent pending) (Figure 1).

**Figure 1.**
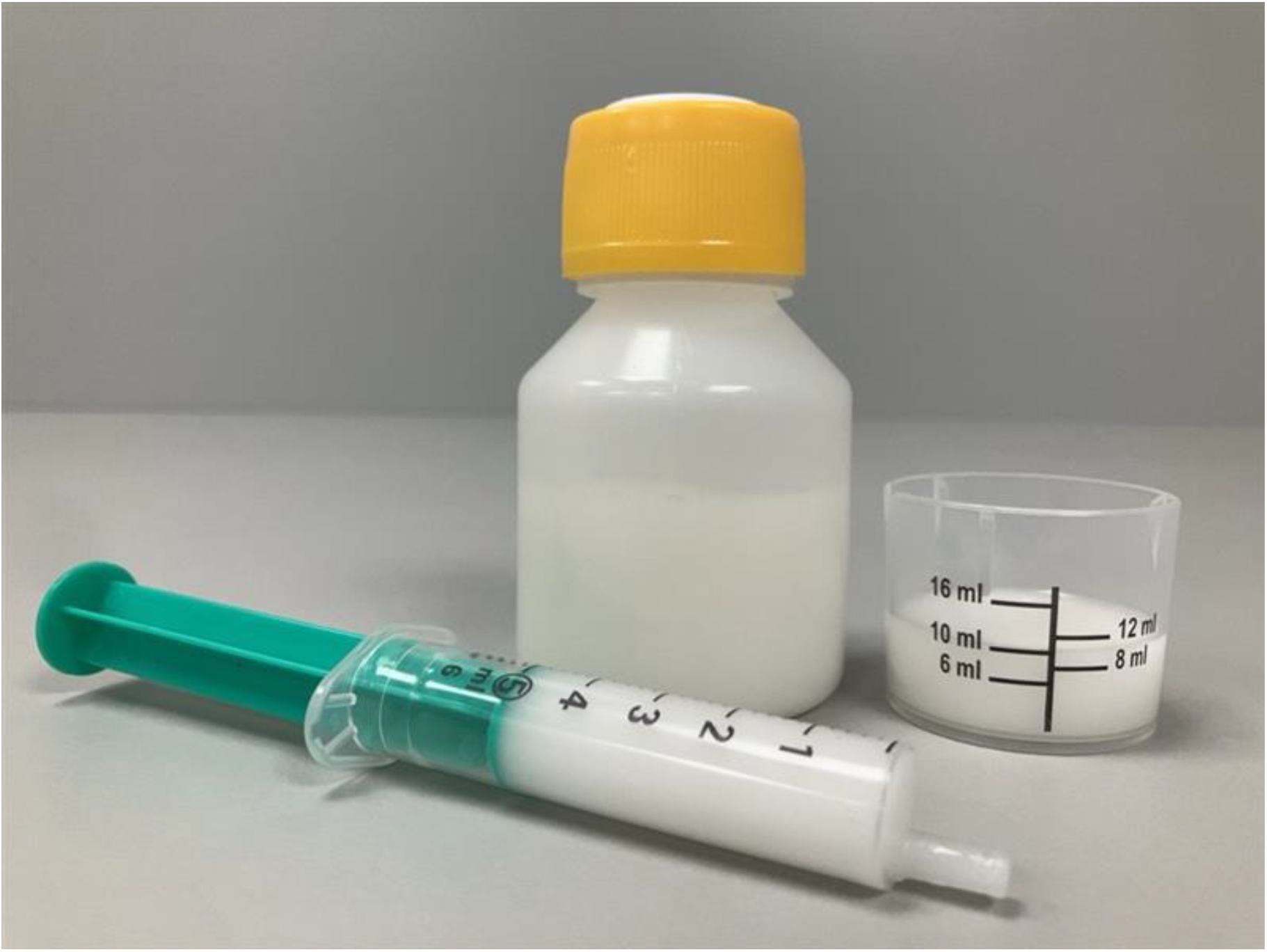
Italfarmaco Quetiapine oral suspension and its presentation with flexible dosing devices.

Italfarmaco quetiapine oral suspension 25 mg/ml (Italfarmaco S.A. Batch n. D21/01 exp date: May 2023) and Seroquel® 25 mg immediate-release film-coated tablets (AstraZeneca Batch SB038, exp date May 2024) were used as Test and Reference products, respectively.

A single-dose, open-label, randomized, two-sequence, two-treatment, two-period crossover pivotal study was conducted in 43 healthy male and non-pregnant female subjects under fasting conditions to compare the pharmacokinetic performance of the quetiapine oral suspension versus immediate release film-coated tablets. Each volunteer received a single 25 mg dose of quetiapine, administered either as an oral suspension or as an immediate-release film-coated tablet, with an appropriate washout between treatments.

In each study period, 17 venous blood samples were collected. The first blood sample was collected prior to drug administration while the others were scheduled to 0:10; 0:20; 0:30; 0:45; 1:00; 1:15; 1:30; 2:00; 2:30; 3:00; 4:00; 6:00; 8:00; 10:00; 12:00 and 24:00 hours:minutes post-dose.

Plasma concentrations of quetiapine were measured using a validated liquid chromatography with tandem mass spectrometry (LC/MS/MS) analytical method. Non-compartmental analysis with Phoenix^®^ WinNonlin^®^ 8.3 was used to derive pharmacokinetic parameters, including maximum observed plasma concentration (C_max_), time to reach C_max_ (t_max_), area under the curve from time zero to the last quantifiable concentration (AUC_0–t_), total exposure (AUC_0–∞_), residual area or percentage of extrapolated part of AUC_0-∞_ (%AUC_extrap_), apparent terminal elimination rate constant (λ_z_), and elimination half-life (t_1/2_). The median difference t_max_ was obtained by calculating the difference for each subject and finding the median of those differences. Study sample analyses were performed by analysts blinded to the dosing randomization scheme.

SAS^®^ 9.4 and Phoenix^®^ WinNonlin^®^ 8.3 were used for statistical analyses. For the pharmacokinetic parameters of interest, an analysis of variance (ANOVA) model was performed.

The terms included were Sequence, Subject nested within Sequence, Period and Formulation. These terms were used as fixed effects and were evaluated at the 5% significance level.

C_max_ and AUC_0–t_ of quetiapine were the primary pharmacokinetic parameters for the bioequivalence assessment. Bioequivalence was inferred if the 90% confidence interval (CI) for the Test-to-Reference Geometric least squares Mean ratio (GMR) of the ln-transformed parameters C_max_ and AUC_0–t_ were both within the 80.00% to 125.00% acceptance interval.

Safety was evaluated through the assessment of adverse events (AEs), 12-lead electrocardiogram (ECG), vital signs, and clinical laboratory tests. For evaluation of safety descriptive, statistics were performed based on the available data only.

The study was approved by following applicable regulations.

## RESULTS

A total of 76 subjects were screened for participation. Of these, 43 healthy volunteers were enrolled in the study and admitted for randomization, including 25 non-pregnant females and 18 males. All 43 subjects completed both study periods and were included in the Bioequivalence Analysis Population. The volunteers had an average age of 34 years (range 21–55), a mean height of 1.68 m (range 1.50–1.89), a mean weight of 70.4 kg (range 47.6–95.2) and a mean BMI of 24.7 kg/m^2^ (range 19.1–29.5) (see Table 2 for demographic data).

**Table 2:** descriptive statistics of demographic data.

| **Demography** | **Parameter** | **Bioequivalence Analysis**<br>**Population**<br>**(n=43)** |
| --- | --- | --- |
| Sex, n (%) | Male | 18 (41.9%) |
|  | Female | 25 (58.1%) |
| Race, n (%) | Black or African American | 5 (11.6%) |
|  | Multiple | 4 (9.3%) |
|  | White | 34 (79.1%) |
| Age (years) | Mean | 34 |
|  | SD | 8.8 |
|  | Median | 32 |
|  | Minimum | 21 |
|  | Maximum | 55 |
| Height (cm) | Mean | 168 |
|  | SD | 8.8 |
|  | Median | 169 |
|  | Minimum | 150 |
|  | Maximum | 189 |
| Weight (kg) | Mean | 70.4 |
|  | SD | 11.9 |
|  | Median | 71.4 |
|  | Minimum | 47.6 |
|  | Maximum | 95.2 |
| Body Mass Index (kg/m<sup>2</sup>) | Mean | 24.7 |
|  | SD | 3.0 |
|  | Median | 24.7 |
|  | Minimum | 19.1 |
|  | Maximum | 29.5 |

n – Number of Subjects; SD – Standard Deviation

### Efficacy

The PK performance is summarized in the Table 3 which displays the results in C_max_, t_max_, AUC from time zero to the last quantifiable concentration (AUC_0–t_), total exposure (AUC_0–∞_), residual area or percentage of extrapolated part of AUC_0-∞_ (%AUC_extrap_), apparent terminal elimination rate constant (λz), and elimination half-life (t_1/2_).

**Table 3:**
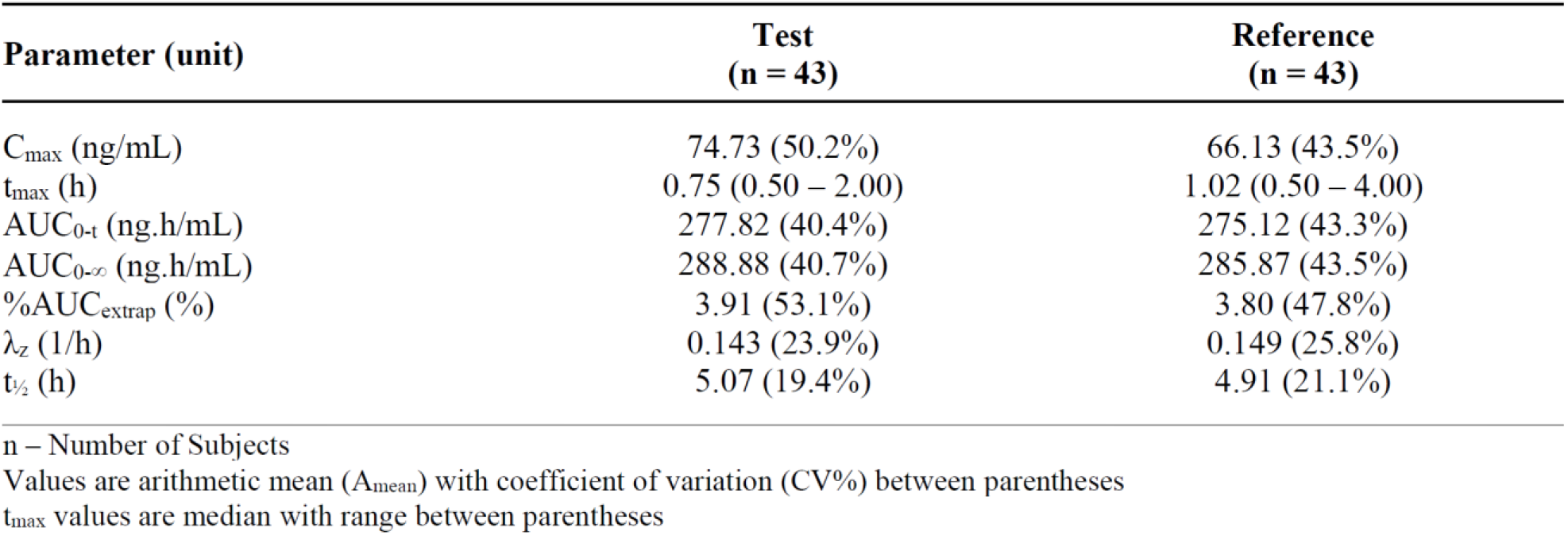
summary statistics of the pharmacokinetic parameters following administration of Italfarmaco quetiapine oral suspension (Test) and film-coated tablet (Reference)

To evaluate bioequivalence between the suspension and the tablet formulations, the Geometric least square Mean Ratio (GMR) of the Test to Reference was calculated for C_max_ and for AUC_0–t_, along with the corresponding 90% confidence intervals (CI). C_max_ ratio was 111.10 (90% CI: 101.25–121.92) and AUC_0–t_ ratio was 101.72 (90% CI: 95.70–108.12) (see Table 4).

**Table 4:** total Intra-Subject Coefficient of Variation (ISCV%) and bioequivalence evaluation by Test-to-Reference Geometric Least Square Means Ratio and corresponding 90% Confidence Intervals for C_max_ and AUC_0-t_.

| Parameter | Total<br>ISCV% | <u>Geometric LSmeans</u> |  | Test-to-Reference<br>GMR (%) | 90% CI |
| --- | --- | --- | --- | --- | --- |
|  |  | Test | Reference |  |  |
| $C_{\max}$ | 26.0 | 67.59 | 60.84 | 111.10 | 101.25 – 121.92 |
| $AUC_{0-t}$ | 16.9 | 255.73 | 251.40 | 101.72 | 95.70 – 108.12 |

Geometric LSmeans values are given in ng/mL for $C_{\max}$ and ng.h/mL for AUC

These results fall within the accepted bioequivalence range of 80–125%, demonstrating that the Test product, Italfarmaco quetiapine 25 mg/ml oral suspension, is bioequivalent to the Reference product, quetiapine 25 mg immediate-release film-coated tablet, after a single dose administration under fasting conditions, in accordance with regulatory guidelines. This demonstrates that the oral suspension can be used as a direct substitute for the tablet at the same dosage, as the equivalent C_max_ and AUC_0–t_ indicates comparable systemic exposure and, consequently, similar expected therapeutic efficacy.

A significant difference was found in t_max_: quetiapine oral suspension reached peak plasma concentration at a median of 0.75 hours (min 0.50 – max 2.00), while immediate-release tablets took 1.02 hours (min 0.50 – max 4.00). The median difference was −0.5 h [90% CI: −0.625 to −0.365; p < 0.001], meaning the oral suspension peaked 30 minutes earlier than the tablet form.

In line with this, quetiapine administered as an oral suspension appeared to be rapidly absorbed, with detectable plasma levels at 10 minutes post-administration and a significant average increase in concentration observed within 20 minutes. In contrast, quetiapine administered as tablet exhibited a slower absorption profile during the early time points (Figure 2).

**Figure 2.**
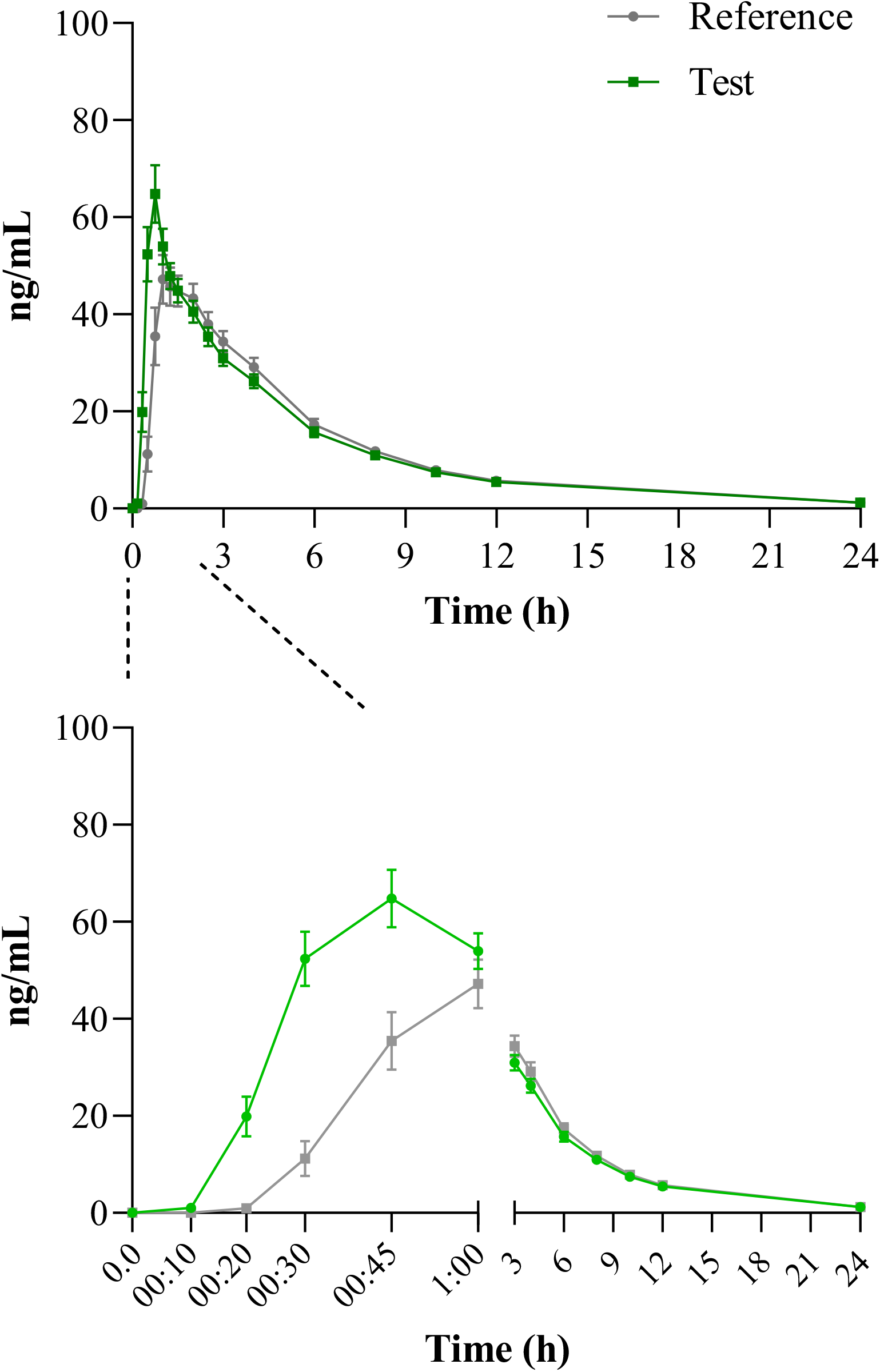
plasma concentration *versus* time profiles following administration of Test and Reference product in linear scale (Upper panel); and overall PK performance of Test and Reference product, zoomed view of the early time points of the curve (Lower panel). Each point represents average and standard error of the mean (SEM)

### Safety

Both Test and Reference products were similarly well tolerated in this study. Out of the 43 subjects who received the investigational products, 38 reported a total of 92 treatment-emergent adverse events (TEAEs).

In the Test group, 34 subjects (79%) experienced 44 TEAEs, of which 34 were considered drug related. The most common TEAE was somnolence, reported by 29 subjects (67%). In the Reference group, 33 subjects (77%) reported 48 TEAEs, with 43 deemed drug related. Somnolence was also the most frequently reported TEAE, occurring in 32 subjects (74%).

All TEAEs were of mild (66) or moderate (26) intensity. No serious adverse events (SAEs) or study discontinuations due to TEAEs were reported.

## DISCUSSION

Managing psychiatric disorders is challenging due to factors like age, comorbidities, polypharmacy, and cognitive or functional limitations that affect treatment adherence and effectiveness.

Treatment of schizophrenia spectrum disorders and bipolar disorders requires a tight control of treatment with respect to side effects, adherence challenges, and patient preferences; this control is crucial for treatment success [4-5].

Among second-generation antipsychotics, quetiapine is widely prescribed due to its broad efficacy and favorable tolerability profile. However, it is predominantly available in solid oral forms, which may limit dosing flexibility and pose challenges for patients with specific needs [1].

In this context, the newly developed Italfarmaco quetiapine oral suspension offers a versatile alternative designed to support tailored treatment strategies. Beyond the inherent advantages of its liquid formulation such as ease of administration and improved patient intake, we observed that this newly developed suspension achieves peak therapeutic plasma concentrations faster, being detectable in plasma within 10 minutes after ingestion.

This earlier absorption profile is consistent with data previously reported for liquid antipsychotic formulations compared with solid forms [1] and an opportunity to achieve a faster onset of action.

The reported time to reach peak plasma concentration for the immediate-release formulation is typically 1–2 hours [8]. Consistently, the immediate-release tablet evaluated in this study reached peak concentration approximately 1 hour after administration.

This formulation demonstrates clinically meaningful value, particularly due to its potential for rapid onset of action. Such pharmacokinetic characteristics are especially relevant in acute clinical scenarios -including insomnia, anxiety, and psychomotor agitation-where timely symptom control is critical. Consequently, this product may offer therapeutic advantages in situations requiring fast symptom relief and prompt clinical efficacy [9].

Although immediate- and extended-release formulations are available [10], having the flexibility to choose the most appropriate medication option remains a critical need for these patients and clinical settings.

The bioequivalence study was performed according to good clinical practice (GCP) standards. The variability values observed among the subjects and reported in Table 4 (Intra-Subject Coefficient of Variation, or ISCV% C_max_ of 26% and AUC_0–t_ 16.9%) are aligned with previous literature reports of 22-28% for C_max_ and 17-18% for AUC_0–t_ [11-12], guaranteeing the representativity of the results obtained.

Other quetiapine suspension formulations have been developed, some at different concentrations; however, published data on their clinical application are still limited. Recent reports have described its real-world use in settings such as Alzheimer’s disease and geriatric populations [13].

To our knowledge, this is the first PK study comparing a quetiapine suspension with its corresponding tablet formulation. These findings highlight the significant role of formulation innovation in advancing more patient-centered approaches to antipsychotic therapy.

## CONCLUSIONS

This study demonstrates that the innovative quetiapine 25 mg/ml oral suspension formulation is bioequivalent to the 25 mg standard immediate-release tablet formulation. However, quetiapine administered as a suspension is readily absorbed, with detectable plasma concentrations 10 minutes after administration. Compared to the tablet form, the suspension reduces 30 minutes the time to reach peak plasma concentration, suggesting a faster onset of action and making it a potentially more suitable option for acute clinical scenarios. This newly developed formulation facilitates administration, dosing flexibility, and patient intake, particularly in patients where factors such as age, comorbidities, polypharmacy, and functional impairments may compromise treatment adherence and efficacy. This is particularly useful for patients requiring low doses or experiencing swallowing difficulties, limited manual dexterity, or cognitive impairment. These features provide clinicians with a valuable alternative to solid oral dosage forms, supporting individualized treatment strategies and enhancing usability in the context of antipsychotic therapy.

## Data Availability

AVAILABILITY OF DATA AND MATERIAL
Data supporting the results reported in the article are available in the corresponding reports and databases, and on request from the authors and/or reviewers.

## SUPPORTING INFORMATION

### FUNDING

This study was funded by ITF Research Pharma, S.L.U., Madrid, Spain.

### AVAILABILITY OF DATA AND MATERIAL

Data supporting the results reported in the article are available in the corresponding reports and databases, and on request from the authors and/or reviewers.

### ETHICS APPROVAL

This study was approved by Portuguese Ethics Committee for Clinical Research (code FG/IFG/2023/1912/20230070) and by INFARMED (DECISION NO. 115/VPCD/2023) and was performed in accordance with the ethical standards as laid down in the 1964 Declaration of Helsinki and its later amendments or comparable ethical standards.

### AUTHOR CONTRIBUTIONS

All authors whose names appear on the submission made substantial contributions to the conception or design of the work; or the acquisition, analysis, or interpretation of data; drafted the work or revised it critically for important intellectual content; read and approved the final version of the manuscript; and agreed to be accountable for all aspects of the work in ensuring that questions related to the accuracy or integrity of any part of the work are appropriately investigated and resolved.

